# Informing People-Centered Target Product Profiles for TB Diagnostics: A Multi-Country Qualitative Study

**DOI:** 10.1101/2025.07.11.25331385

**Authors:** Maria del Mar Castro, Hien Le, Kingsley Manoj Kumar, Elisabeth Mamani-Mategula, Annet Nakaweesa, Victoria Dalay, Kinari Shah, Rouxjeane Venter, Devasahayam J. Christopher, Charles Yu, Grant Theron, William Worodria, Payam Nahid, Adithya Cattamanchi, Claudia M. Denkinger, Andrew D. Kerkhoff, Nora West, Ha Phan

## Abstract

**Background:** Timely and accurate tuberculosis (TB) diagnosis remains a key challenge in high-burden settings. The World Health Organization (WHO) has developed Target Product Profiles (TPPs) to guide diagnostic development, which have largely reflected the perspectives of experts, with limited input from people affected by TB. This qualitative study explored preferences and experiences to inform people-centered TB diagnostic strategies.

**Methods:** We conducted 75 semi-structured interviews with adults undergoing TB evaluation at outpatient clinics in India, the Philippines, South Africa, Uganda, and Vietnam. Participants were purposively sampled to ensure diversity in sex, TB status, age and treatment. Thematic analysis was utilized.

**Findings:** Preferences were shaped by five interrelated domains: perceived diagnostic accuracy, sample collection experience, time-to-results, affordability, and testing location. Diagnostic accuracy was consistently prioritized, with many expressing willingness to trade comfort and convenience for more trustworthy results. Sputum and blood were widely trusted despite collection challenges, whereas tongue swabs and urine were easier to provide but perceived as less accurate. Rapid, same-day turnaround was valued for minimizing emotional distress, financial and logistical burdens. Although testing was typically free, indirect costs such as transport and lost income, remained barriers. Hospital-based testing was preferred due to trust in staff and infrastructure, though some acknowledged the appeal of community-based approaches if reliability and privacy were ensured.

**Conclusion:** People seeking TB care prioritize accuracy and trustworthiness, even at the expense of comfort or convenience. These preferences can inform WHO policy updates, especially regarding the adoption of novel sample types and testing strategies, to support uptake and equitable access to novel diagnostics.

## Introduction

Tuberculosis (TB) remains the leading cause of death from a single infectious agent and continues to pose a significant global public health challenge. An estimated 10.8 million people had TB in 2023, of which 8.2 million people were newly diagnosed and notified with TB, with the highest burden in the WHO South-East Asia (45%), African (24%), and Western Pacific (17%) regions [1]. TB is both preventable and curable with timely diagnosis and treatment [2]. Despite advances in diagnostics and treatment, large gaps persist across the TB care cascade, particularly at the stage of diagnosis [3, 4]. Missed or delayed diagnoses remain a major contributor to ongoing transmission, morbidity, and mortality [5]. In high-burden settings, people with TB often encounter barriers at every step of their care journey, from recognizing symptoms and seeking care, to accessing timely and accurate testing, receiving a diagnosis and treatment. Global and national strategies emphasize the need to strengthen diagnostic capacity to eliminate TB and reduce preventable deaths and suffering [6].

Closing these gaps requires a deeper understanding of the experiences and preferences of people affected by TB. To achieve this goal, global health efforts emphasize people-centered models of care [7], and a growing body of work explores what patients themselves value in the TB diagnostic process [8–10]. The World Health Organization (WHO) has developed Target Product Profiles (TPPs) to guide the development of novel TB diagnostics, outlining minimal and optimal criteria for different test’s attributes, such as sensitivity, specificity, cost, time to results, sample type, and ease of use [11]. However, these specifications have largely been informed by the perspectives of researchers, policy-makers, and implementers, while the views, values, and priorities of people affected by TB have not been systematically incorporated.

This qualitative research is part of a broader multi-country project exploring people preferences for novel TB diagnostic tools. As part of this work, our team recently conducted a discrete choice experiment (DCE) to quantify preferences in relation to the WHO TPP attributes for an ideal diagnostic test [10]. Findings from the DCE indicate that diagnostic accuracy, rapid results, and cost-free testing are the most highly valued features among individuals seeking TB testing [10]. Building on these findings, we conducted semi-structured interviews in five high TB burden countries to understand the drivers of these preferences and to explore broader preferences related to the care seeking process. Insights from this qualitative study will inform the design and evaluation of TB diagnostic strategies that are more responsive to patient priorities and support future revisions of the WHO TPP for TB diagnostics.

## Methods

### Study design

This qualitative study was conducted between July 2023 and August 2024. Semi-structured interviews were conducted among adults with symptoms of TB who presented for evaluation at outpatient clinics participating in the R2D2 TB Network [12], where both novel TB tests and standard-of-care diagnostics were being evaluated. The study aimed to explore participants’ perspectives, experiences, and preferences on various aspects of the TB diagnostic process, including pathways to TB testing, sample collection, perceived accuracy and trust in different test types, result notification and turnaround time, cost and preferences for testing settings. Reporting of findings follows the Consolidated Criteria for Reporting Qualitative Research (COREQ) guidelines [13]. The checklist is provided as **Supplementary File 1**.

### Study setting

This study was conducted at outpatient clinics in five countries: India, the Philippines, South Africa, Uganda, and Vietnam. Sites were purposively selected from facilities participating in the R2D2 TB Network, which supports evaluations of novel diagnostic tools. Across sites, a range of care levels was represented, from tertiary referral hospitals to primary health clinics, encompassing urban and peri-urban settings. Detailed site characteristics have been previously described [10].

In brief, in India, participants were recruited from a large tertiary hospital in Vellore and two additional outpatient clinics (one in a nearby district and one in a secondary-level facility in Chittoor, Andhra Pradesh). In the Philippines, recruitment occurred at a hospital-based DOTS clinic in a peri-urban area outside Manila. In South Africa, participants were enrolled from a public clinic in a peri-urban district of Cape Town. In Uganda, interviews were conducted at the outpatient TB clinics of a national referral hospital and a municipal hospital within urban Kampala. In Vietnam, participants were recruited from a lung hospital in urban Hanoi.

All study sites offered routine symptom-based screening for TB. Under standard of care, most facilities relied on sputum-based molecular diagnostics. As part of the R2D2 study, novel diagnostic tests using sample types such as urine, tongue swabs, and blood were also evaluated. Results of these evaluations were not used to diagnose or initiate TB treatment. Facilities had the necessary infrastructure, staffing, and laboratory capacity to implement both standard and experimental diagnostic workflows.

### Study population

Participants were purposively sampled using a maximum variation approach [14]. Sampling aimed to ensure diversity across gender and TB test results (positive, negative, or pending) criteria. We further considered age and treatment status (on treatment vs. not yet initiated) in sampling participants. We sought to include 15 participants per country (75 total) to ensure representation across diverse sites while capturing variation in experiences and preferences. This pragmatic sample size decision reflected the multi-country study design and operational feasibility. We anticipated that thematic saturation across countries would be met with the total sample size, and the study team reviewed memos and transcripts throughout the data collection process to ensure saturation was achieved [15].

### Data collection

A topic guide (**Supplementary file 2**) was developed and reviewed with local teams to ensure appropriateness, cultural relevance, and clarity. Pilot interviews informed minor revisions based on feedback from site teams. Transcripts from the first two participant interviews from each country were translated into English and reviewed centrally to provide feedback and reinforce interviewing techniques prior to following interviews. Interviews were conducted by trained local investigators and study staff, most of whom were female and had varying experience in qualitative research. All data collectors participated in qualitative data collection and standard operating procedure (SOP) training, including review and discussion of the interview guide to promote consistency and rigor.

Semi-structured interviews were held in private spaces within the facilities, typically on the day of TB evaluation, or within two weeks of initial evaluation, if needed. They lasted between 60–90 minutes, were audio recorded, and field notes were documented using debriefing forms. Automated transcription [16] was utilized in India (for English speaking participants), while manual transcription was needed in South Africa, Vietnam, Uganda and Philippines, due to limited performance of the tool. All transcripts were reviewed against the audio by site staff to ensure quality. Revised transcripts were translated into English for analysis, and clarifications were obtained from data collectors through email or teleconference as needed. MMC, EM, and NW led periodic debriefings with each country team to monitor progress, ensure data quality, and discuss emerging insights.

### Data analysis and interpretation

Thematic analysis was used [17], combining inductive and deductive approaches. Two researchers with experience in qualitative research (MDM and EMM) began with independent, close readings of early transcripts from each country to familiarize with the data and inductively generate initial codes. Codes were iteratively discussed and organized through reflexive engagement with the data. To ensure alignment with study aims, we also considered how the evolving codes related to the core domains explored in the interview guide. A preliminary codebook was developed and refined in consultation with NW and the broader research team.

EMM coded the remaining transcripts using ATLAS.ti (www.atlasti.com), iteratively adding new codes until thematic saturation was achieved. Regular meetings with MDM were held to resolve discrepancies, consolidate overlapping codes, and finalize the thematic structure. Upon completion of coding, codes were clustered and refined into candidate themes through iterative team discussions. Themes were then reviewed for coherence, distinctiveness, and interpreted in relation to the study objectives, including the features of the TPP for TB diagnostics [11]. Analytic memos were used to support theme development and interpretation. During analysis, comparisons were made across study countries, sex and TB status to identify commonalities and site-specific perspectives.

### Ethical considerations

Ethical approval for this study was obtained from the local Institutional Review Boards (IRBs) in each participating country, as well as from the University of California, San Francisco, and the Heidelberg University Hospital. All participants provided written informed consent.

## Results

We conducted 75 semi-structured interviews with participants from India, the Philippines, South Africa, Uganda, and Vietnam (**Table 1**). Most participants were male (56% overall), with median age of 35 years (IQR=24-46), and 49% tested positive for TB. HIV status varied by site, with 11% of participants testing HIV-positive, primarily from South Africa.

**Table 1.** Characteristics of study participants.

| Characteristic | Overall<br>N = 75<sup>1</sup> | India<br>N = 15<sup>1</sup> | Philippines<br>N = 15<sup>1</sup> | South Africa<br>N = 15<sup>1</sup> | Uganda<br>N = 15<sup>1</sup> | Vietnam<br>N = 15<sup>1</sup> |
| --- | --- | --- | --- | --- | --- | --- |
| **Age** | 35 (24-46) | 32 (22-42) | 40 (28-54) | 33 (20-45) | 30 (22-36) | 45 (26-59) |
| **Sex** |  |  |  |  |  |  |
| Female | 33 (44%) | 7 (47%) | 7 (47%) | 9 (60%) | 6 (40%) | 4 (27%) |
| Male | 42 (56%) | 8 (53%) | 8 (53%) | 6 (40%) | 9 (60%) | 11 (73%) |
| **TB test result** |  |  |  |  |  |  |
| Negative | 38 (51%) | 7 (47%) | 8 (53%) | 8 (53%) | 8 (53%) | 7 (47%) |
| Positive | 37 (49%) | 8 (53%) | 7 (47%) | 7 (47%) | 7 (47%) | 8 (53%) |
| **TB treatment status** |  |  |  |  |  |  |
| Started | 33 (44%) | 8 (53%) | 7 (47%) | 7 (47%) | 7 (47%) | 4 (27%) |
| Not started / not applicable | 42 (56%) | 7 (47%) | 8 (53%) | 8 (53%) | 8 (53%) | 11 (73%) |
| **HIV status** |  |  |  |  |  |  |
| Negative | 50 (67%) | 14 (93%) | 0 (0%) | 9 (60%) | 13 (87%) | 14 (93%) |
| Positive | 8 (11%) | 0 (0%) | 0 (0%) | 6 (40%) | 2 (13%) | 0 (0%) |
| Unknown | 17 (23%) | 1 (6.7%) | 15 (100%) | 0 (0%) | 0 (0%) | 1 (6.7%) |

<sup>1</sup>Median (Q1-Q3); n (%)

### Overview of preferences and tradeoffs across TPP attributes

Participants’ accounts revealed that preferences for TB diagnostic tests were shaped by a combination of interrelated attributes, including perceived accuracy, time-to-result, cost and affordability, testing setting, and the comfort or ease of sample collection (**Figure 1**). These attributes were often considered in relation to one another, rather than in isolation, with participants describing the trade-offs they were willing to make based on their circumstances and priorities. Among these, the balance between the perceived accuracy and the ease or comfort to produce a sample was widely discussed. Additionally, familiarity with certain sample types and with the health system, as well as trust in the testing process influenced how participants interpreted and valued test attributes.

**Figure 1.**
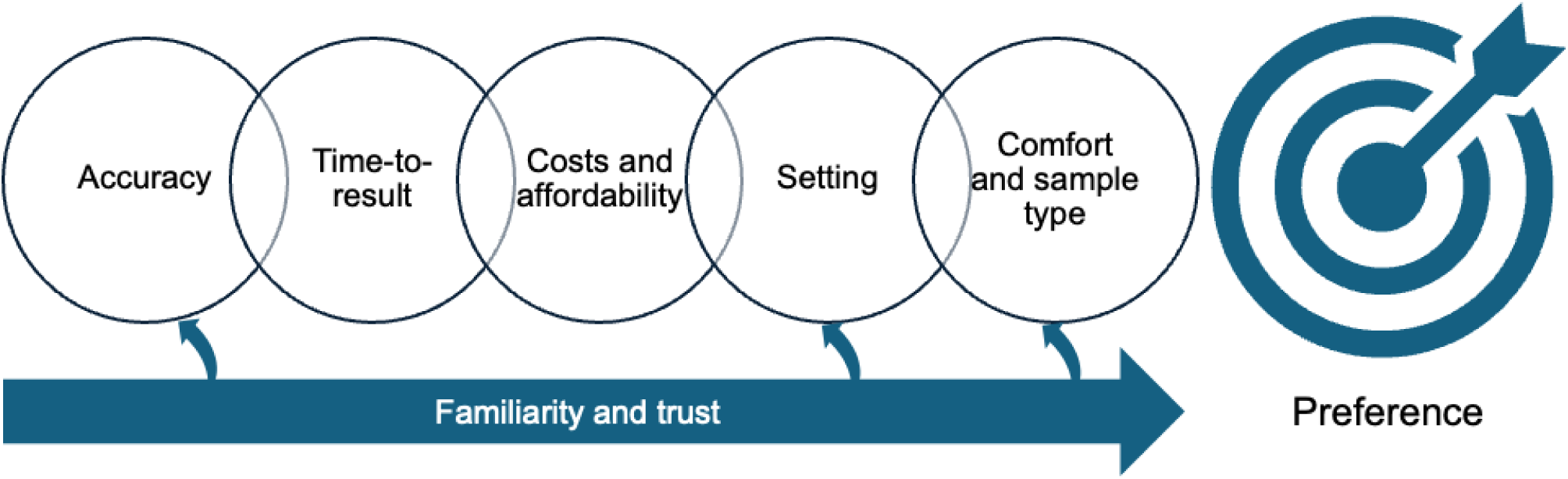
Attributes influencing diagnostic preferences.

### Context: pathways to TB testing and experiences with sample types

Participants described a range of symptoms that led them to seek TB testing, most commonly persistent cough, chest pain, fatigue, weight loss, and shortness of breath. Some sought initial care at pharmacies or local clinics and turned to hospitals when symptoms did not improve. In South Africa, Uganda, and the Philippines, participants often went directly to hospital-based TB services. In India and Vietnam, participants were more likely to be referred from local facilities to specialized hospitals. Frequently the decision to seek testing was influenced by advice from family or friends, or concern about contact with someone known to have TB.

*“I realized that I was slowly losing my life. I said why am I still going to these clinics and pharmacies. Let me go to the hospital and get tested, and they will see what is happening with this cough that is hurting from the inside.” (Female, TB Negative, Uganda)*

Participants described giving a range of biological samples for TB testing, including sputum, tongue swabs, blood and urine. Although not a sample, participants also mentioned chest X-rays as part of the diagnostic process. Sputum was the most frequently collected sample across countries and was often described as the most difficult to produce. Many reported struggling with unproductive cough, physical discomfort, and needing to walk or exercise to induce sputum.

*“They made me exercise and I was in pain. Getting sputum was a challenge for me. Even when I got it, it was very little because my cough was not productive. That is where I found challenges. When I got some little sputum, I brought it to the health worker, and they told me to get more. There were four bottles, but I had failed to gather it, so they made me exercise. But I still failed. I was only able to get a little.” (Female, TB Positive, Uganda)*

Others received support through nebulization.

*“My experience is not difficult. Nebulizer was put in my mouth and I tried to produce more sputum and after induction I was able to give sample.” (Male, TB Negative, India)*

Tongue swabs were commonly viewed as easy and comfortable. Participants described the procedure as quick, causing minimal discomfort, and familiar due to previous experiences with COVID-19 testing.

*“Best is tongue swab, I just need to sit, and they will touch my tongue and take the sample.” (Female, TB Negative, India)*

However, some expressed concerns about sample contamination from food or saliva. Some men reported feeling embarrassed by tongue swab collection because of having to show or open their mouth. while some women described discomfort, itching and nausea during sample collection.

“*The worry I had was that the doctor was going to see the inside of my mouth or the doctor smelling the bad odor from my mouth. So I was thinking about that. It went on well.” (Male, TB Negative, Uganda)*

Blood samples were generally accepted and viewed as routine. Both female and male participants described discomfort with venipuncture, pain, or fear of needles. They described concerns about the volume of blood taken or the lack of explanation given about it. Notably, some participants emphasized the minimal effort or personal agency involved in blood and tongue swab collection, contrasting it with sputum or urine collection, which required active engagement (e.g., walking, or attempting to produce a sample).

“*For giving blood sample I just need to sit, for giving urine, I need to wait for when it comes, similarly for giving sputum I need to try and wait for a while*.” (Female participant, TB Negative, India)

Urine collection was generally seen as easy and familiar, especially among women with experience from testing for pregnancy or urinary tract infections. However, some female participants found it unhygienic, and others mentioned difficulty urinating on demand. Some male participants also expressed disgust on collecting the sample. Chest X-rays were often perceived as a routine part of the diagnostic work-up. They were generally described as straightforward and acceptable, although it was not always available in the same facility where initial testing took place.

Regarding instructions received prior to testing, most participants described them as clear, and that staff provided support throughout the process. Some noted improvements compared to past visits, where guidance was limited or unclear. Participants often credited the presence of research staff for helping them find the right testing areas. However, some said they were not told why samples were being collected or what the testing involved, indicating gaps in communication.

*“The instructions were easy because the nurse explained it thoroughly to me and that made it easy to follow them.” (Female, TB negative, South Africa)*

### Perceived accuracy and trust in sample types

Many participant narratives of the diagnostic testing process highlighted a tradeoff between trust in a test’s ability to accurately diagnose TB and the comfort or ease of producing a sample for that test.

Sputum and blood were typically viewed as more accurate but collecting them was often uncomfortable or physically demanding. In contrast, tongue swabs were easier to provide but not always seen as accurate. This tension shaped participants’ preferences, with some prioritizing ease while most noted a willingness to endure discomfort to ensure a trustworthy result. (**Figure 2**)

**Figure 2.**
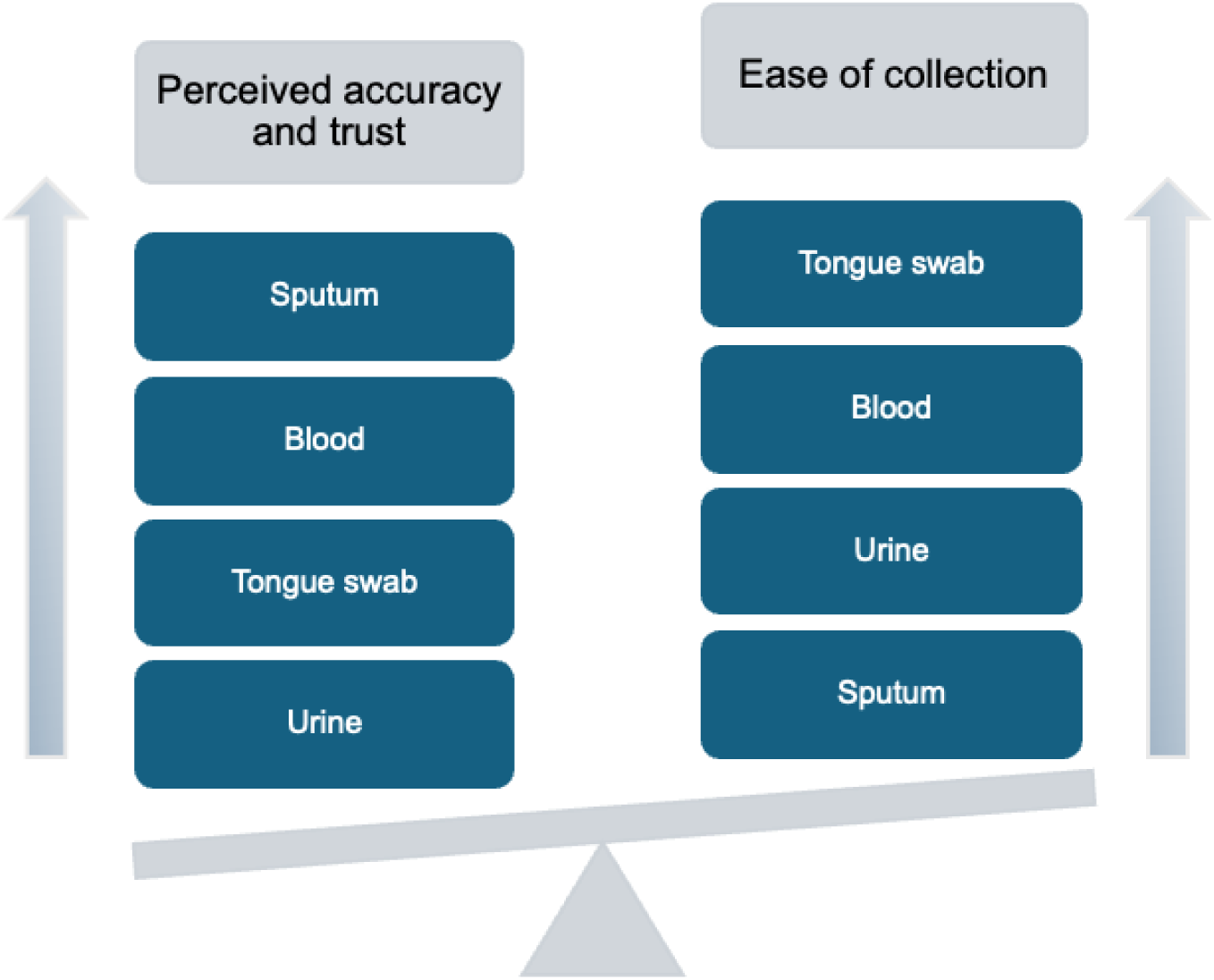
Balance between perceived accuracy and ease of collection regarding sample preferences. Arrows indicate increasing rank (from lowest at the bottom to highest at the top), and the tilted scale illustrates that perceived accuracy and trust often outweighed ease of collection in shaping preferences.

*“If the tongue swab result is as accurate [as the sputum specimen test result] then I would prefer the tongue swab.” (Male, TB Negative, Vietnam)*

Regarding specific sample types, sputum was most frequently identified as the sample that best confirmed TB, largely because it was understood to come directly from the lungs. Many participants associated it with long-standing practices in TB diagnosis and described it as the most trusted method.

*“Because it [sputum] really comes from the lungs, it’s like it’s the actual phlegm from the lungs, so its accuracy is really high, like out of 10, it might be 9 accurate. It really comes from the lungs.” (Female, TB Negative, Philippines)*

Blood samples were also seen as reliable by some participants, who mentioned that blood is used to diagnose other diseases. Some felt that blood tests were simpler and less prone to error than sputum, which they believed could be affected by poor sample quality.

*“I would have liked the method of blood withdrawal. I choose it because I think we are used to blood tests it is a normal thing, and we know it is the one that brings results.” (Male, TB Positive, Uganda)*

Tongue swabs were viewed as accurate by some participants, particularly when sputum was difficult to produce or blood was avoided. However, others were unsure about the reliability of tongue swabs, especially if the collection process was rushed or if there were food particles or saliva in the mouth.

*“I feel tongue swab testing would be easier, as I told you earlier, I faced difficulty in giving sputum.” (Male, TB positive, India)*.

*“Well, the advantage [of tongue swab] is that sometimes it’s really easy. You just open your mouth, stick out your tongue, rub the swab around, and that’s it. The disadvantage is that… I’m not really familiar with it [laughs], but it’s still close to the lungs. (Male, TB positive, Philippines)*.

Chest X-rays were mentioned less often but were sometimes considered highly accurate because they provide a visual view of the lungs. A few participants said they preferred X-rays for their ability to “see inside” and detect lung damage. In contrast, urine samples were rarely considered reliable for TB diagnosis. Some participants said they did not understand how urine could be used to detect a lung disease and expressed uncertainty about its accuracy.

*“The advantage of urine in tuberculosis, maybe, ma’am, it’s an advantage for the patient, it’s easy to extract urine, but in terms of tuberculosis testing, it seems a bit far off, ma’am.” (Female, TB Negative, Philippines)*

When prompted about the possibility of false positive or negative results, participants reflected on the emotional and practical implications of inaccurate TB tests. False-positive diagnoses often provoked strong emotional reactions, including frustration, sadness, and anger. Many expressed they would feel betrayed by the health system, especially when treatment had already begun. Concerns centered on unnecessary exposure to side effects, the physical toll of treatment, as well as the financial (e.g., loss of work) and social consequences of being labeled as having TB.

*“In general, I would be very sad and actually regretful. It is difficult because I trusted the hospital when I came to the hospital. I would lose my faith in the hospital in the future (Male, TB Negative, Vietnam)*

False-negative results generated a different but equally intense response. Participants described feelings of fear, helplessness, and loss of confidence in the diagnostic process, especially when symptoms persisted without explanation. Several expressed a desire to seek care at alternative facilities, particularly larger or more specialized hospitals, to ensure a more accurate diagnosis.

*“I will go for another examination to find out why I’m still coughing and having chest pain, even though I was told I don’t have the disease. I will go to a different facility to get checked again.” (Male, TB Negative, Vietnam)*

In contrast, some participants said they were more likely to return to a familiar facility for follow-up or a second opinion, even if the test result was unexpected. Trust in the institution outweighed doubts about individual tests, particularly when prior care had been perceived as effective.

### Time to results and notification

Participants across all study sites expressed a clear preference for receiving TB test results on the same day, with many citing an ideal timeframe of receiving results on the same day by noon. Faster turnaround was valued for convenience and reduced emotional distress, including the possibility of prompt treatment, reduced disruption to work and household obligations like school drop-offs. The desire for timely results was often driven by concerns about prolonged uncertainty, fear of contracting TB from others at the facility, or potentially transmitting the disease to others (e.g., relatives) if diagnosis was delayed.

*“What I would have loved them to change is the time that results take to come out because if someone is waiting for their results, they think it would take a short time, they already have a calculation of some sort saying maybe when I am done, I will go and work and then they find that they have been caught up with time, and time has gone and they can’t go to work anymore, so they count that day wasted because they have not earned.” (Female, TB Negative, Uganda)*

Despite this preference, participants reported a range of experiences with turnaround times. In Uganda, the Philippines, and Vietnam, patients frequently arrived at health facilities early in the morning (around 7:30 AM) and received their results by mid-afternoon. Although the process was generally completed within the same day, participants still described the wait as emotionally taxing and logistically burdensome. In South Africa, results were typically available within 24 hours, with some participants receiving results via SMS or phone calls. In contrast, the India site had the longest time-to-results, with some participants reporting having to stay near the hospital or return for results often after two or more days. These logistical challenges, especially for those traveling long distances, imposed financial strain and created additional uncertainty.

*“Maybe there will be no one to look after the children, I can’t leave my father alone with the children.” (Female, TB Negative, Philippines)*

Interviewees emphasized their willingness to return for follow-up visits and result collection. Many framed this commitment as a matter of life and health, noting they would rearrange personal schedules to ensure they received their results. However, barriers like work-related conflicts, and the strain of balancing childcare and home responsibilities for women, were mentioned. Transport expenses, although not universally prohibitive, were mentioned by many participants as a source of stress, particularly when multiple visits were required. Participants suggested reducing turnaround time by increasing staff, improving scheduling, and streamlining result delivery.

*“I would have returned because it is upon my life. Personally, I would have returned the day I am told even if it takes my time or sabotages my plans I would return on that day that I would have been told.” (Male, TB Positive, Uganda)*

### Preferences for testing settings: Hospital vs community

Most participants preferred hospital-based TB testing. Hospitals were viewed as more reliable, better equipped, and staffed by trained professionals. Many participants said they felt more confident receiving a diagnosis in a facility with specialized tools, such as X-rays and laboratory services. Hospitals were also seen as places where additional health concerns could be addressed during the same visit. Although reaching a hospital sometimes involved long travel or time off work, participants generally considered these burdens acceptable in exchange for receiving a thorough and accurate evaluation.

*“Hospital for me because at least in the hospital, everything is available. It’s complete, and at least they can explain things properly to you… all the necessary treatments are available.” (Female, TB Negative, Philippines)*

Some participants mentioned that community- or home-based testing was more convenient, especially for those living far from hospitals or with limited transportation. Participants worried that neighbors might see them being tested at home, leading to unwanted attention or assumptions about their health. Others expressed doubt that community-based services had the proper equipment or trained staff to carry out TB testing.

*“Actually, I think I agree that those locations are convenient, but I cannot trust them for this kind of disease. If I have taken time to get tested, so I will go to a reputable facility, at least, I have to go to a nearby hospital. I cannot trust those places.” (Male, TB Negative, Vietnam)*

However, concerns about lack of specialized care, privacy, stigma, and the reliability of testing outside hospital settings were common.

*“I trust here at the hospital because there are specialized doctors here. At our health center, they have schedules like “Oh, today the doctor is not here,” and the doctor goes to other barangays.” (Female, TB Negative, Philippines)*

There were also concerns about the quality of specimens collected outside of hospitals, including delays in transport and exposure to heat. Some participants felt that these issues could affect the accuracy of the results and lead to repeat testing.

### Financial burden and affordability

Across country sites, participants consistently reported that the core components of TB diagnostic services, including consultations, sample collection and test results, were provided free of charge. Many described this zero-fee structure as essential to accessing care, noting that financial barriers would have otherwise prevented them from seeking testing. Participants, mainly from the Uganda site, frequently emphasized that even a small fee would deter care-seeking, especially for individuals with irregular or unstable incomes. Free testing was viewed as a critical enabler of early detection and treatment.

*“It would not have been easy for me because my income is not consistent and I take a long time to make an income, but because it was free of charge, it was easy for me and that made me happy.” (Male, TB Negative, Uganda)*

Despite the absence of direct charges for TB testing, many participants reported indirect costs associated with diagnosis. These included expenses for transportation, food, accommodation, and in some cases, additional tests or medications not covered under standard services. Such costs were particularly burdensome for participants who had to return multiple times or wait long hours for results.

*“I know that the testing here is free, so there’s no need to spend any money. But we paid for our transportation, which was just 24 pesos round trip.” (Female, TB Negative, Philippines)*

In Vietnam, unlike other sites, participants noted a reliance on national health insurance to offset costs. Insurance typically covered a large proportion of diagnostic expenses, and participants paid the remaining (approx. 5%). This system was viewed as fair and not prohibitive, with several participants expressing trust in the hospital billing process.

*“When I stay at the hospital, I will receive health insurance benefits which I am entitled to.” (Female, TB negative, Vietnam)*

## Discussion

This qualitative study highlights how patient preferences for TB diagnostic tests are shaped by a combination of perceived accuracy, sample collection comfort, time to results, testing location and affordability. Participants consistently emphasized the importance of tests that are trusted, minimally burdensome, and rapidly delivered, with many expressing a willingness to endure physical discomfort or logistical barriers in exchange for a more reliable result. However, others preferred easier tests if they perceived them to be reasonably accurate, revealing a nuanced balance between diagnostic trust and user experience.

A DCE conducted by our team [10] showed that diagnostic accuracy, short time to result, and free of charge were the most influential attributes shaping decision-making. In addition, people were willing to trade lower accuracy or a small fee for faster results. The qualitative findings contextualized these tradeoffs, illustrating how trust, prior experiences, and comfort shaped participants’ preferences. While the WHO TPP for diagnostics considers time to result delivery as an implementation issue [11], our findings show that people seeking TB care perceived it as an intrinsic feature of test quality. Previous studies reported the willingness of people to stay longer at the facility for a same-day result [18, 19], and the DCE showed overall preference for rapid results [10]. The emotional, logistical and financial burden, including the loss of work due to long waiting times partly explain these tradeoffs, for example, a small fee may be lower than the opportunity cost of waiting for hours at the facility. Long waiting times and administrative complexities have also been described previously as a source of diagnostic delays [20].

While the TPP focuses on cost per test, people seeking TB diagnosis prioritized affordability in a broader sense, including indirect expenses and opportunity costs. Pre-diagnosis expenditures can account for a large portion of TB-related costs [21–23], with some studies estimating them as 50% of the costs of drug-susceptible TB [24]. These financial burdens are known to disproportionately affect women and people on the lowest income brackets [24]. In our study, even small user fees were seen as a potential deterrent to testing, among those with unstable incomes, emphasizing the critical role of cost-free services and other social protection measures [25] in ensuring access. Preference for facility-based testing was driven by the perceived quality and reputation of diagnostic services, with the value of adequate diagnostics outweighing the effort to access them. While hospital-based testing was generally trusted and preferred, community-based testing was viewed as more convenient. This emphasizes the continued efforts to improve the quality of TB services, which is a priority in the global End TB strategy [26, 27]. However, this study involved participants who reached the health facilities and were typically symptomatic -people who may be more likely to prioritize diagnostic quality over convenience when actively worried about their health - which may underrepresent the perspectives of those minimally symptomatic or utilizing community services.

The importance of accuracy aligns with the WHO TPPs, which defines technical specifications for analytic sensitivity [11], and is consistent with previous qualitative and mixed-methods studies that identified accuracy as the most valued test attribute [10, 28]. This expanded to the selection of testing settings and type of samples. In our study, preferences for sample types were shaped by perceived accuracy above factors like comfort and ease of collection. This is the case of sputum-based testing, which was preferred due to its familiarity and perceived link to the lungs [10, 19]; non-sputum sampling was also valued by participants, despite acknowledged limitations [18, 29, 30]. The mixed views on sample types highlight the need for multiple options, especially for those unable to produce sputum [31, 32]. Recent publications suggest that more accessible, lower-cost tests with slightly lower sensitivity may achieve greater equity and uptake [33], as well as increase diagnostic yield [34]. Realizing these benefits will require effective communication, and recognition of how perceived accuracy shapes sample type preferences, during the rollout of novel, non-sputum-based tests.

Our findings also revealed people’s concerns about false-positive diagnoses, including emotional distress, loss of trust in the healthcare system, and the burdens of unnecessary treatment. Recent studies have shown higher mortality rates among individuals clinically diagnosed with TB compared to those with bacteriologically confirmed disease[35], partly due to missed opportunities to treat underlying conditions. Additional consequences include exposure to side effects of unnecessary treatment, delays in appropriate care for non-TB illnesses [36, 37], and the financial burden and the stigma associated with a TB diagnosis [38]. These insights underscore the need for diagnostic tools that balance sensitivity and specificity, minimizing the harms from misclassification.

This study has several strengths. It draws on a large and diverse sample from five high TB burden countries, enhancing the geographic and contextual relevance of the findings. The inclusion of participants with varying TB diagnosis and treatment status and balanced gender representation enabled a nuanced understanding of diagnostic preferences. Rigorous quality assurance procedures such as piloting, interviewer training, and regular debriefings with country teams [39], supported the consistency and contextual appropriateness of data collection. However, several limitations should be acknowledged. The analysis relied on translated transcripts, which may have led to some loss of nuance or misinterpretation [40], despite close collaboration with local teams and translation or review being done by bilingual team members involved in data collection. The study only included individuals who were able and willing to access health facilities, potentially underrepresenting the views of people facing significant access barriers [3, 9], which is particularly relevant when interpreting preferences for testing location. Since participants were all adults, we were not able to explore preferences among adolescents, children, or their caregivers, who may prioritize test attributes differently. Additionally, some of the scenarios explored were hypothetical, and stated preferences may differ from actual behavior in real-world settings [41]. Future work should examine the views of those excluded from formal care pathways and their stated preferences.

In conclusion, this study highlights the need to align TB diagnostic tools with the lived realities and preferences of people seeking care. Participants prioritized tests that are accurate, affordable, easy to complete, and deliver rapid results; characteristics they perceived as essential to test quality. These findings suggest that people-centered perspectives can inform future revisions of target product profiles (TPPs) and diagnostic evaluation frameworks. Integrating the priorities of people affected by TB into test design, selection, and rollout will be critical to ensuring both equitable access and meaningful uptake of novel diagnostics.

## Data Availability

All data produced in the present study are available upon reasonable request to the authors.

## Acknowledgments

The authors would like to thank all persons undergoing tuberculosis testing who participated in the study. We also extend our gratitude to the staff and administrative teams at the participating research sites for their valuable support. The work at the Christian Medical College, Vellore site was done under the auspices of TB RePORT India consortium, supported by the Department of biotechnology, India and NIAID, USA.

