## Supplementary file 1 for "Informing People-Centered Target Product Profiles for TB Diagnostics: A Multi-Country Qualitative Study"

#### Contents

Supplementary file 1. COREQ checklist....2

Supplementary file 2. Topic guide....4

#### Supplementary file 1. COREQ checklist

### COREQ (COnsolidated criteria for REporting Qualitative research) Checklist

| Topic | Item No. | Guide Questions/Description | Reported on [explanation added if needed] |
|---|---|---|---|
| **Domain 1: Research team and reflexivity** | | | |
| *Personal characteristics* | | | |
| Interviewer/facilitator | 1 | Which author/s conducted the interview or focus group? | Trained site researchers for each country conducted all the interviews. |
| Credentials | 2 | What were the researcher's credentials? E.g. PhD, MD | The research team included members with MD, MSc, MPH, and PhD qualifications. |
| Occupation | 3 | What was their occupation at the time of the study? | Described in the Methods section, under "Data collection". |
| Gender | 4 | Was the researcher male or female? | |
| Experience and training | 5 | What experience or training did the researcher have? | |
| *Relationship with participants* | | | |
| Relationship established | 6 | Was a relationship established prior to study commencement? | No prior relationship existed between interviewers and participants. Interviewers introduced themselves at the start of data collection and responded to participants' questions related to the study. |
| Participant knowledge of the interviewer | 7 | What did the participants know about the researcher? e.g. personal goals, reasons for doing the research | Interviewers introduced themselves and their institutional affiliations. The study objectives and the purpose of the research were explained during the consent process. |
| Interviewer characteristics | 8 | What characteristics were reported about the inter viewer/facilitator? e.g. Bias, assumptions, reasons and interests in the research topic | Interviewers were nationals of the respective study countries, with experience and research interest in TB diagnostics. |
| **Domain 2: Study design** | | | |
| *Theoretical framework* | | | |
| Methodological orientation and Theory | 9 | What methodological orientation was stated to underpin the study? e.g. grounded theory, discourse analysis, ethnography, phenomenology, content analysis | Described in the Methods section, under "Study design." |
| *Participant selection* | | | |
| Sampling | 10 | How were participants selected? e.g. purposive, convenience, consecutive, snowball | Described in the Methods section, "Study population" |
| Method of approach | 11 | How were participants approached? e.g. face-to-face, telephone, mail, email | Described in the Methods section, under "Data collection". |
| Sample size | 12 | How many participants were in the study? | Methods section, under "Study population", Table 1 |
| Non-participation | 13 | How many people refused to participate or dropped out? Reasons? | |
| *Setting* | | | |
| Setting of data collection | 14 | Where was the data collected? e.g. home, clinic, workplace | Described in the Methods section, under "Data collection". |

| | | | |
|---|---|---|---|
| Presence of nonparticipants | 15 | Was anyone else present besides the participants and researchers? | |
| Description of sample | 16 | What are the important characteristics of the sample? e.g. demographic data, date | Table 1 |
| **Topic** | **Item No.** | **Guide Questions/Description** | **Reported on Page No.** |
| *Data collection* | | | |
| Interview guide | 17 | Were questions, prompts, guides provided by the authors? Was it pilot tested? | Appendix 2. |
| Repeat interviews | 18 | Were repeat inter views carried out? If yes, how many? | N/A (no repeated interviews) |
| Audio/visual recording | 19 | Did the research use audio or visual recording to collect the data? | Described in the Methods section, under “Data collection”. |
| Field notes | 20 | Were field notes made during and/or after the inter view or focus group? | |
| Duration | 21 | What was the duration of the inter views or focus group? | |
| Data saturation | 22 | Was data saturation discussed? | Described in the Methods section, under “Study population” and “Data collection” |
| Transcripts returned | 23 | Were transcripts returned to participants for comment and/or correction? | |
| **Domain 3: analysis and findings** | | | |
| *Data analysis* | | | |
| Number of data coders | 24 | How many data coders coded the data? | Described in the Methods section, under “Data analysis and interpretation”. |
| Description of the coding tree | 25 | Did authors provide a description of the coding tree? | |
| Derivation of themes | 26 | Were themes identified in advance or derived from the data? | |
| Software | 27 | What software, if applicable, was used to manage the data? | ATLAS.ti (www.atlasti.com) |
| Participant checking | 28 | Did participants provide feedback on the findings? | We did not seek participant feedback on the findings. |
| *Reporting* | | | |
| Quotations presented | 29 | Were participant quotations presented to illustrate the themes/findings? Was each quotation identified? e.g. participant number | Quotations are presented throughout the Results section. Each each quotation is identified by basic participant characteristics (sex, TB status and country) |
| Data and findings consistent | 30 | Was there consistency between the data presented and the findings? | Described in the Results section; themes are identified as subheadings. |
| Clarity of major themes | 31 | Were major themes clearly presented in the findings? | |
| Clarity of minor themes | 32 | Is there a description of diverse cases or discussion of minor themes? | Described in the Results section. |

Developed from: Tong A, Sainsbury P, Craig J. Consolidated criteria for reporting qualitative research (COREQ): a 32-item checklist for interviews and focus groups. *International Journal for Quality in Health Care*. 2007. Volume 19, Number 6: pp. 349 – 357

#### Supplementary file 2. Topic guide

Topic guide for semi-structured interviews with patients or people seeking diagnosis for TB
Version 2.0 21 June 2023

*Information for the researcher:*
***Set-up of the interview****: Ensure that the location is comfortable for the participant. Ideally, it should not have background noise, ensure that the interview is not interrupted, and provide the needed comfort and privacy. Provide refreshments if available.*

***Information and consent****: Ask the participant if consent for participating in the study was already obtained as part of the DCE survey. If not, provide Information and Consent Form (ICF) to the participant, inform on reason of interview, study question and recording of interview. Invite the participant to clarify outstanding questions and obtain written consent prior to starting the interview. Check that consent was given and ensure that the participant is offered a signed copy of the ICF.*

*The interview aims to identify relevant values, preferences, and outcomes of the diagnostic process for TB patients in high TB burden countries and explore their relationship to TB diagnostic test preferences.*

*This is an interview guide and not a semi-structured questionnaire. Therefore, your approach with every participant should be quasi-conversational. You should also be attentive to covering the domains identified, but not every interview is expected to cover all domains. You are the one to decide when to follow a participant's lead if it provides useful new data for the project. The probes listed below are suggestions of questions within a given domain. You do not have to ask all the questions, and some may be redundant.*

*Be sure to collect the following information and enter it on the summary sheet at the beginning of the interview.*

- Interview Date:
- Interview Location:
- Interviewer Name:
- Participant ID:

| **Topics and subitems** | **Prompt** |
|---|---|
| **Introduction and warm-up** | |
| Introductions | Good morning/afternoon. Thank you for taking the time to participate in this interview. My name is ______.<br><br>**<u>Purpose of this interview:</u>**<br>We are gathering information from people with presumptive TB, to understand their perspectives on using new tests that could potentially allow timely diagnosis and more rapid treatment initiation. We would like to know your experience, preferences and difficulties while seeking TB diagnosis, and its process which you have just experienced.<br><br>There is no right or wrong answer. Please feel free to express your views during this interview. Your responses will be kept private and only myself and the research team will read them. Your answers will not be linked to your name and will not affect the treatment or care you will receive.<br><br>The interview will last approximately ***1 – 1.5 hour***. We will be recording the interview so that we can go back and review what was said later. We will also be taking notes to help us along with the recording. If there are any questions that you feel uncomfortable answering, we can skip to the next question or stop the interview. |

| | |
|---|---|
| | Do you have any questions so far? |
| Warm-up (Opening question) | To start, tell me about yourself, and the reasons that motivated you to seek testing for TB? |
| **Experiences, preferences and challenges with diagnostic testing for TB** | |
| Diagnostic Process | Can you please describe your experience getting tested for TB?<br><br>*Alternative phrasing*: Tell me about the day you got tested for TB, from the time you decided to go to the clinic to the time when you went home.<br><br>*Probe: What challenges did you encountered when getting tested for TB?*<br>*Probe: What did you like or dislike when getting tested for TB and why?* |
| Methods/types of tests | How did you feel when the health worker suggested that you be tested for TB?<br><br>*Probe: Please tell me about any worries or concerns you had about being tested for TB.* |
| Sample collection | Please tell me about which sample(s) was required for the test(s) for TB? When I say sample I mean how you were tested and what you gave as part of your TB test. *[Ask for each type of sample]:*<br><br>What were your experiences producing or giving [insert sample type]?<br><br>*Probe: How did you find producing or giving the sample?*<br>*Probe: What made it easy or difficult to produce or give the sample(s)?*<br><br>Please tell me how you felt about the instructions that were given to you by the HCP to collect the sample(s)<br><br>*Probe: Tell me about anything that was easy to understand about the instructions.*<br>*Probe: Tell me about anything that was hard to understand about the instructions.* |
| Preferences regarding samples | Thinking about the different types of samples you might give for a test *[provide examples/descriptions]* (sputum into a cup, urine, tongue swab, or blood). Is there any sample you trust or prefer the most? Please tell me why.<br><br>Based on your experience providing sputum samples, if you could give an alternative sample instead, what would you give?<br><br>*[Ask about Saliva, tongue swab, urine, finger prick]*<br>*Probe: What do you think about tongue swab? [Ask about each of them, saliva, urine, etc.]*<br>*Probe: What advantages/disadvantages might this have compared to sputum?*<br><br>Please tell me for diagnosing TB, which type of sample do you feel more comfortable giving? (Sputum, urine, blood, swabs) and why? |

| | |
|---|---|
| Distance | Can you describe your travel experience to the place where you were tested for TB, including the time it took to reach the facility.<br><br>*Probe: What means of transportation did you use?*<br>*Probe: How far do you have to travel to visit the TB clinic?*<br>*Probe: Transportation method? Challenges?*<br><br>If the test result needed a few days to be confirmed, would you have to come back or stay in the area to wait? Please tell me about why.<br><br>If you need to stay, how do you manage (place and costs) to stay in the city/town while waiting for the results? |
| Preferences regarding alternatives to minimize issues related to distance | If you could pick any place to get tested for TB that is not at a health facility, what would it be? And why?<br><br>*Probe: Is there any place where you would NOT want to be tested for TB? What makes you say that?*<br><br>*Probe: Tell me your thoughts on getting tested at home.*<br><br>*Probe: Tell me your thoughts on getting tested somewhere in the community.*<br><br>*Probe: Having samples being transported for processing? (e.g., concerns about their sample being transported away or waiting longer for the results)* |
| Support during the diagnostic process | When you had to seek testing for TB, tell me about any kind of help you got from family, friends, neighbors, or others?<br><br>*Probe: Who? What helped you most?*<br>*Probe: What help did you need that you did not get?*<br><br>Please tell me about any concerns you had about family, friends, neighbors or other people knowing that you were seeking testing for TB?<br><br>*Probe: Did concerns about TB stigma impact who you sought support from?* |
| Time to results and making sense of results | Please tell me about how long each step of your experience took from the time you left your house to when you got the clinic, to when you got your test result<br><br>Can you tell me your thoughts about the time it took to receive results?<br><br>*Probe: Does the time you waited for the results matter for you?*<br>*Probe: What did you do in the meantime (waiting time)?*<br><br>Please tell me about how you were notified of the result of your TB test?<br><br>*Probe: Where did you go or what did you do to get your results?*<br><br>How were the results of your tests explained to you?<br><br>[*If positive*] What does a positive result mean to you? |

| | |
|---|---|
| Availability of the tests [If applicable] | When you were tested for TB, tell me about payment for the test?<br><br>How did having to pay for the test factor into your decision to get the test or to seek healthcare when you felt sick?<br><br>Was there a time when you go for a test and you were told the test is bad/had error or it/machine is not working? Please describe. |
| Follow-up | Are there things that prevented you from coming back to receive your results or initiate treatment, if any?<br><br>*Probe: Distance, children at home, being too sick to travel*<br><br>Were you asked to tell members of your household to get tested or checked for TB? (Contact testing). If yes, can you tell me your experience?<br><br>*Probe: Please tell me about your experience asking members of your household to get tested or checked for TB.*<br>*Probe: What made it easy or difficult to tell members of your household to get tested?* |
| Adverse events or negative consequences<br><br>(If not explored in the diagnostic process) | Please tell me about any problems during or after the tests for TB were done that were the result of taking the test.<br><br>*Probe: Please tell me about any experiences you had with discrimination or negative treatment as a result of seeking testing or get a TB diagnosis.*<br><br>*Probe: Please tell me about any concerns you had related to TB stigma when seeking testing for TB.* |
| False negatives | There is no test that is always able to detect TB on the first time of testing. If you were to receive a negative result on a TB test, what would you think?<br><br>Please tell me about what you would do if your symptoms didn't get better but the test for TB says you do not have TB?<br><br>*Probe: Would you try to get another test for TB – why or why not?*<br>*Probe: Would you seek care or treatment somewhere else? If yes, where would you go?* |
| False positives | Although rare, there is a small chance that some tests show a patient has TB when they really do not. If this rare situation happened to you, and you learned that your positive result (meaning you are sick) was actually incorrect, how would you feel?<br><br>*Probe: How would it affect your life? What would you do and why?* |
| Expectations | After going through the process of being tested or test and diagnosed, what would you change for people undergoing the testing process for TB?<br><br>*Probe: What would you change about the sampling, location, or duration of the testing for TB?*<br><br>Please describe for me any |
| Closing/conclusion | We have reached the end of our interview. Do you have anything else to add that we have not discussed already? |

| |
|---|
| Thank you for taking the time for this interview |

*At the end of the interview, write a summary of any practical details (e.g., setting, etc.) that you see fit and offer your reflections on what you have just reviewed. You should consider situating your reflective notes directly at the end of the summary as this may work best for you rather than across each domain.*
